# New Zealand Emergency Department COVID-19 Preparedness Survey

**DOI:** 10.1101/2021.04.06.21253178

**Authors:** Michael J. Howard, Charlotte Chambers, Nicholas M. Mohr

**Author notes:** Corresponding Author: Michael J. Howard MD PhD.

## Abstract

**Objective:** Our objective was to assess the level of COVID-19 preparedness of emergency departments (EDs) in Aotearoa New Zealand (NZ) through the views of emergency medicine specialists working in district health boards around the country. Given the limited experience NZ hospitals have had with SARS-CoV-2, a comparison of current local practice with recent literature from other countries identifying known weaknesses may help prevent future healthcare worker infections in NZ.

**Methods:** We conducted a cross-sectional survey of NZ emergency specialists in November 2020 to evaluate preparedness of engineering, administrative policy, and personal protective equipment (PPE) use.

**Results:** A total of 137 surveys were completed (32% response rate). More than 10% of emergency specialists surveyed reported no access to negative pressure rooms. N95 fit testing had not been performed in 15 (12%) of respondents. Most specialists (77%) work in EDs that cohort COVID-19 patients, about one-third (34%) do not use spotters during PPE doffing, and most (87%) do not have required space for physical distancing in non-patient areas. Initial PPE training, simulations and segregating patients were widespread but appear to be waning with persistent low SARS-CoV-2 prevalence. PPE shortages were not identified in NZ EDs, yet 13% of consultants do not plan to use respirators during aerosol generating procedures on COVID-19 patients. Available treatments including non-invasive ventilation and high-flow nasal cannula were common.

**Conclusions:** New Zealand emergency specialists identified significant gaps in COVID-19 preparedness, and they have a unique opportunity to translate lessons from other locations into local action. These data provide insight into weaknesses in hospital engineering, policy, and PPE practice in advance of future SARS-CoV-2 endemic transmission.

**Strengths and limitations of this study:**

- Survey responses specifically identified existing breakdowns in engineering, administrative policy and personal protective equipment in New Zealand emergency departments, potentially increasing healthcare worker nosocomial infection risk upon reintroduction of SARS-CoV-2
- Survey included emergency specialists from all 20 of New Zealand’s district health boards but the electronic convenience sample may not be representative of all ED consultants in NZ
- Some survey questions asked respondents to recall experiences or project how they would practice if they were caring for a COVID-19 patient and those motivated to respond may feel they have more or less access to protective policies and equipment than non-respondents

## INTRODUCTION

The Aotearoa New Zealand (NZ) healthcare system was as unprepared for the coronavirus disease 2019 (COVID-19) pandemic as many nations, yet NZ has successfully eliminated severe acute respiratory syndrome coronavirus 2 (SARS-CoV-2).(1, 2) Geographic isolation and a lag in cases behind other nations in mid-March 2020 gave NZ time to assess preparedness using several epidemiologic models of pandemic mortality and healthcare demand.(3, 4) The decision to implement aggressive public health infection elimination practices hinged on NZ’s ability to rapidly and effectively close its borders thus limiting COVID-19 impact to approximately 2600 cases and 26 deaths.(5, 6) As a result, NZ’s Emergency Departments (EDs) have had little experience caring for COVID-19 patients and disparate efforts towards infection control preparedness may leave heath care workers (HCWs) vulnerable to endemic SARS-CoV-2 transmission.(7-10)

The Hierarchy of Control offers an algorithm to assess preparedness of a health system, scalable to departmental, hospital and nationwide recommendations.(10-12) Once elimination is established but eradication remains impossible there must be appropriate resources to institute and sustain substitution of the threat (typically by vaccination or other therapies). Even as vaccine-based immune protection expands there are still uncertainties requiring multiple controls to prevent transmission of SARS-CoV-2. Questions about viral variants that evade host immune responses, vaccine safety and efficacy in vulnerable groups (ie. young children, immunocompromised, elderly), and the impact of vaccine hesitancy indicate we will need to maintain layers of protection for some time into the future.(13) In addition to vaccination, pandemic ED response should continue focus on proven non-pharmaceutical interventions such as engineering (often through changes in ED physical layout, ventilation and bed allocation), administrative policy (infection prevention and control, workflow changes, training, resources), and transmission-based PPE. These practices demand equity, and the failure to maintain high-quality controls has resulted in healthcare worker (HCW) infections, disability and death.(10, 14-16).

The July-August 2020 outbreak in Melbourne, Victoria, Australia revealed deficiencies in hospital level infection prevention and control (IPC) in a health system comparable to that of Aotearoa New Zealand.(17, 18). Unfortunately, this outbreak in long-term care facilities and subsequent nosocomial spread in tertiary hospitals resulted in significant SARS-CoV-2 infections in HCWs. The Australian response affords insight into system preparedness improvements to adopt in other health systems.(10, 19, 20) The New Zealand Emergency Department COVID-19 (NZEDC19) Preparedness Survey of emergency consultants was designed to identify and address weaknesses in local NZ emergency department policy, engineering, and PPE to provide proactive recommendations for system improvement.

## METHODS

This study was a cross-sectional web-based assessment of COVID-19 pandemic preparedness of EDs in NZ via survey of ED senior medical officers (ED SMOs) from the EDs of all NZ District Health Boards (DHBs).

### Questionnaire Design

A 27-item questionnaire was framed around the hierarchy of control model with questions on engineering (negative flow isolation rooms, shared/cohorted patient areas, segregated patient flow, physical distancing), administrative controls (policies for rostering, training, simulations, treatments, and breaches), and personal protective equipment (supply, fit testing, use and re-use).(10, 12, 21) A series of Likert scale questions evaluated individual stress, wellness and risk assessment. Questions were adapted from a published survey of preparedness in intensive care units (ICUs) of Australasia and the prospective COVID Evaluation of Risk in Emergency Departments (COVERED) Project in the U.S.(22, 23) The questionnaire was validated using established survey methodology, after several rounds of consensus building process between ED, microbiology and infectious disease specialists.(24)

### Survey Distribution

The survey was distributed by email to 422 members of the Association of Salaried Medical Specialists (ASMS) identified as having emergency medicine as their designated department of work using Survey Monkey (San Mateo CA, USA) between 26 October 2020 and 23 November 2020. Two e-mail reminders were sent. Participation was voluntary. The study was considered exempt from the institutional review board by the NZ Health and Disabilities Ethics Committees.

### Data Analysis

The data analysis was primarily descriptive and reported as percentages of valid responses. Diverging stacked Likert scales are used to display emergency specialist opinion results. The survey and raw data are included as a supplemental appendix.

### Patient and Public Involvement

Patients or members of the public were not involved in the design, or conduct, or reporting, or dissemination plans of this research.

## RESULTS

One-hundred thirty-seven surveys were completed (32% response rate). All (100%) of 20 NZ DHBs were represented by at least 2 returned surveys. Nine (6.6%) respondents did not identify a DHB.

Engineering: The majority of respondents have access to negative flow or negative pressure patient care rooms (Table 1). Most (115, 83%) report 4 or fewer such rooms in their ED, but 14 (12 %) ED specialists reported no access to negative flow rooms for COVID-19 patient care. Most respondents worked in EDs that have some beds separated only by curtains with shared air circulation where patients may be cohorted. Most (n=126, 92%) surveyed emergency consultants work in EDs which can create physical separation of care areas for high (HIS) index of suspicion patients segregated from those for presumed low index of suspicion (LIS) patients. Emergency consultants from multiple DHBs commented that ED segregated flow or “streaming” can be changed with COVID-19 prevalence and alert level.

**Table 1:** Summary table of select NZEDC19 Preparedness Survey answers

| **Control** | **Specific hierarchy of control question** | **N** | **%** |
| --- | --- | --- | --- |
| **Engineering** | Have negative flow/pressure rooms in ED | 123 | 88% |
|  | Have cohorted beds in ED | 99 | 77% |
|  | Segregated COVID/non-covid patients in ED | 126 | 92% |
|  | Rostered to see both COVID/non-COVID as needed | 70 | 60% |
|  | Unable to meet physical distance requirements at office | 94 | 70% |
|  | Unable to meet physical distance requirements at workstation | 118 | 87% |
|  | Unable to meet physical distance requirements at break rooms | 92 | 71% |
| **Policy** | Intubate LIS patient in negative pressure | 6 | 4% |
|  | Intubate HIS patient in negative pressure | 88 | 64% |
|  | Dedicated intubation teams ICU/anaesthesia | 57 | 47% |
|  | Intubation of HIS/COVID with video laryngoscopy | 98 | 71% |
|  | Use HFNC for hypoxic COVID-19 patients | 53 | 50% |
|  | Use NIV for hypoxic COVID-19 patients | 101 | 86% |
|  | Use NIV with in-line expiration viral filter | 19 | 16% |
|  | No PPE training | 3 | 2% |
|  | PPE group training in-person with observed practice | 66 | 37% |
|  | PPE individual training in-person with observed practice | 40 | 23% |
|  | Simulation training of intubation in COVID-19 patient | 93 | 70% |
|  | Simulation training of NIV in COVID-19 patient | 61 | 46% |
|  | Simulation training of self-proning in COVID-19 patient | 17 | 13% |
|  | Not monitored during donning PPE | 39 | 30% |
|  | Not monitored during doffing PPE | 44 | 34% |
| **PPE** | Not N95 fit tested by time of this survey | 15 | 12% |
|  | Fit tested by qualitative method (odour or taste) | 82 | 60% |
|  | Fit tested by quantitative method (machine sampling) | 41 | 30% |
|  | Wear N95 for HIS/COVID patient not receiving AGP | 61 | 48% |
|  | Wear N95 or PAPR for AGP of HIS/COVID patient | 110 | 87% |
|  | N95 masks unavailable | 6 | 6% |
|  | Re-use N95 masks without sterilization | 12 | 11% |
|  | Re-use N95 masks after sterilization | 3 | 3% |
|  | Elastomeric respirators unavailable | 63 | 66% |
|  | PAPRs unavailable | 79 | 82% |

Most respondents did not feel they could meet minimum physical distancing requirements for patient care in their workplace and disagree or strongly disagree that physical distancing is possible at offices (n=94, 70%), workstations (n=118, 87%), and break rooms (n=92, 71%) (Figure 1).

**Figure 1:**
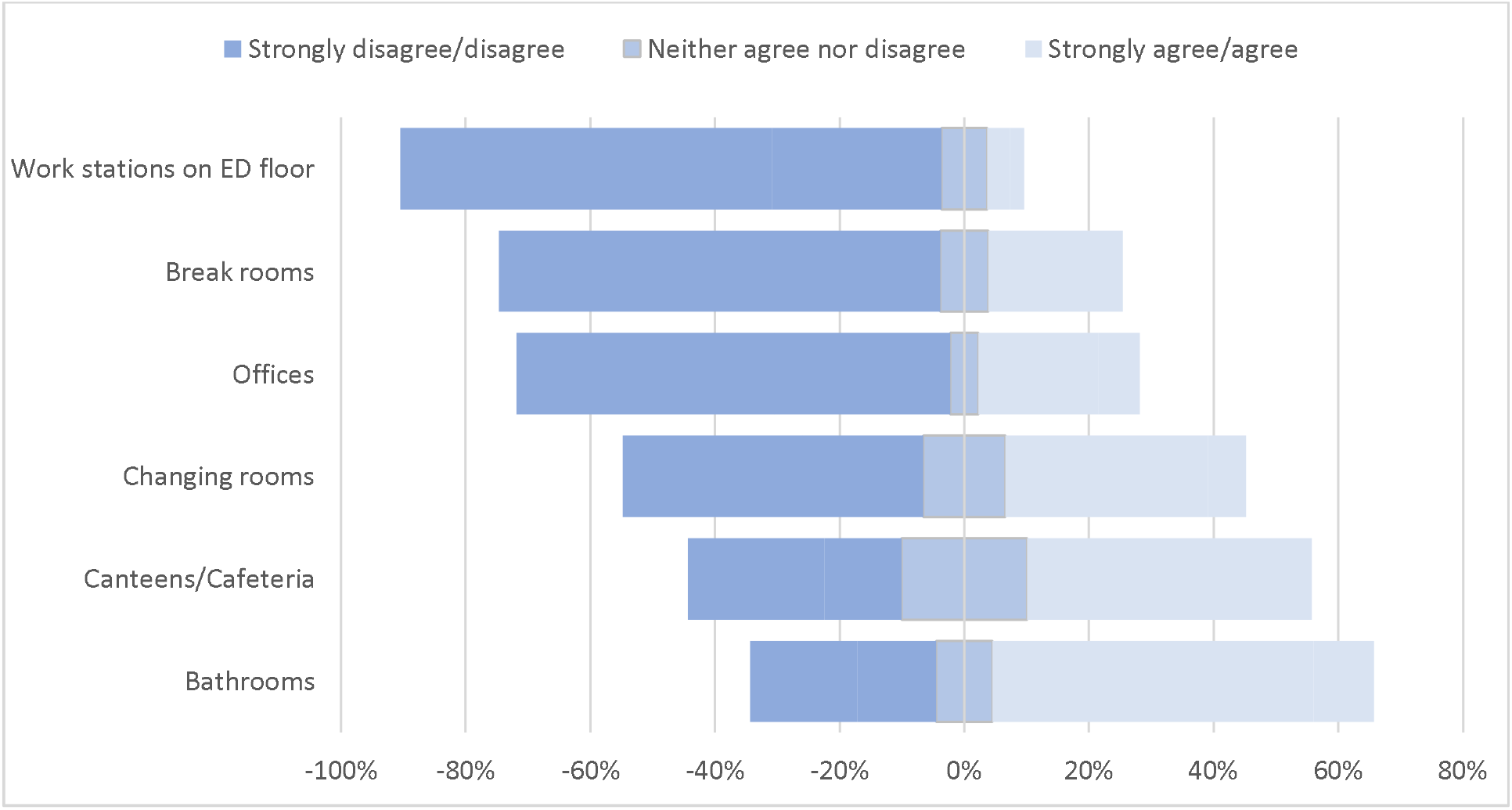
Do you feel able to meet minimum physical distancing requirements in the non-clinical staff areas of your ED?

Administrative controls: Rostering ED consultants into either strictly “COVID” or “non-COVID” teams is not common and the majority (n=70, 60%) see these patient populations during their work period. Almost all (98%) of NZ ED consultants report having training for proper transmission-based PPE use with 60% having had in-person sessions being observed donning and doffing by the instructor. In practice, NZ emergency specialists report donning observation is rarely (18%) mandatory and about a third (30%) do not have an observer present. Only 16% report mandatory observation during removal while a third (34%) are not usually observed doffing PPE. Greater than half the NZ emergency consultant workforce surveyed are not aware of an official breach-of-PPE policy in their hospital ED or breach criteria.(25) Although they report access to showers, approximately 75% of NZ ED SMOs report no policy for showering after a recognized breach, after shifts, or at home.

Simulation training is common in NZ for patient intubation (93, 70%). Less common simulations are performed for non-invasive ventilation (61, 46%) and are rare for patient self-proning (17, 13%).

Greater than half (54%) of specialists report HFNC availability, but 14% would not use this technology at all. Half (55%) of ED specialists say they can utilize NIV (CPAP/BiPAP) but only 16% report using viral expiration filters, a low-cost recommended infection control adaptation. NIV is not used outside negative pressure rooms and only 4% report need to transfer to ICU for this modality. The majority of specialists report wide discretion in their ability to apply NIV to COVID-19 patients and just 15% reserve it only for patients with comorbidities (COPD, CHF, etc.) known to benefit.

Sixty-four percent of consultants would intubate HIS/COVID-19 patients in a negative pressure room. Very few (4%) would intubate patients screened as LIS/non-COVID-19 patients under negative pressure.

The lack of adequate staffing levels during the pandemic is cited as the greatest concern for two-thirds of respondents. Having adequate PPE and adequate testing capacity if a future wave of COVID-19 occurred in NZ were less concerning for respondents (Figure 2).

**Figure 2:**
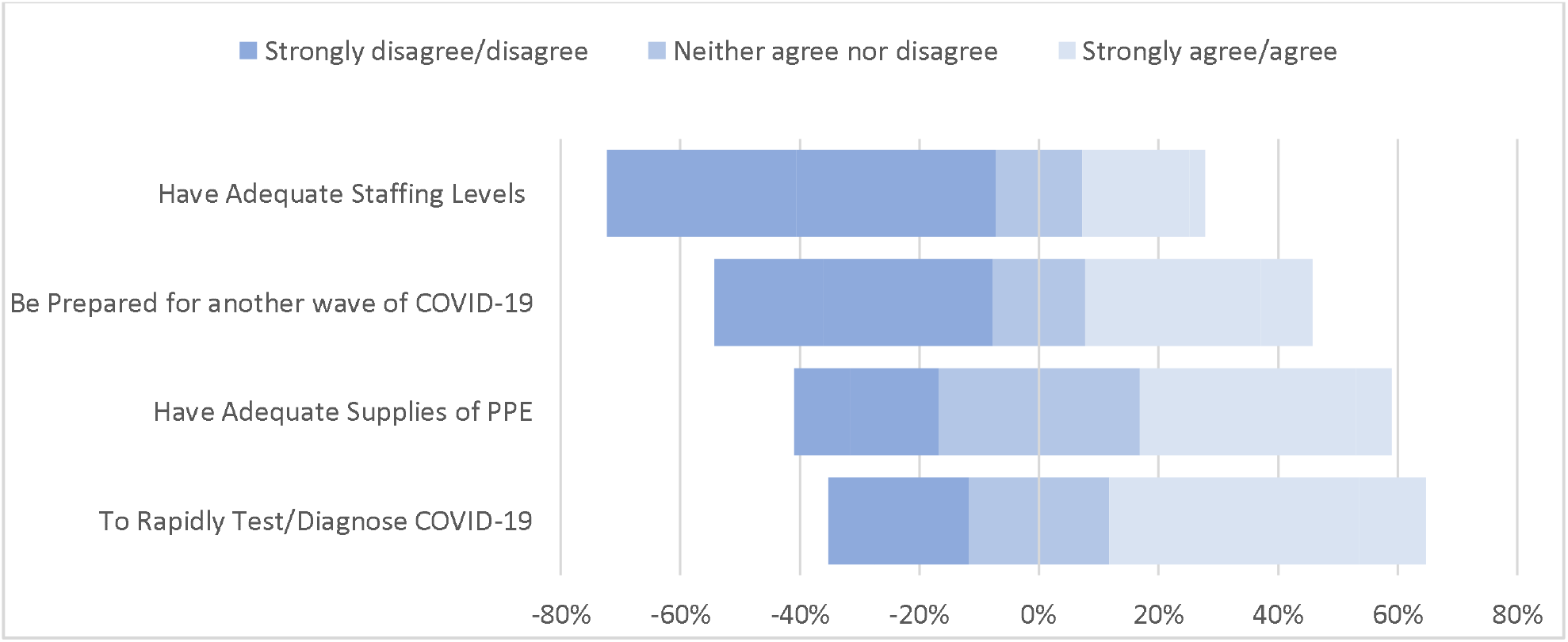
If there were another wave of COVID-19 in NZ, what are your views regarding your ability to do the following:

Personal protective equipment: New Zealand emergency consultants report few shortages of consumable PPE and have had little experience with reusing PPE, except washable face shields and goggles (Table 1). Low reuse of N95 masks either without sterilization (9%) and after sterilization (2%) further supports that respondents felt PPE supplies were adequate. Few respondents reported use of elastomeric respirators (2%) and powered air-purifying respirators (PAPRs) (2%).(26, 27)

Only 89% of respondents had been fit tested for N95 masks at the time of this survey, leaving approximately one in ten of ED consultants surveyed (11%) having not been fit tested by November 2020. Half of these (7/15) were from one hospital.

Best practice for ED consultant use of transmission based PPE was assessed in different clinical scenarios as shown in (Table 2). Only 83% of respondents reported they would use N95 respirators in the context of aerosol generating procedures (AGP), with an additional 4% protected with elastomeric or PAPR. Thirteen percent of respondents would not use a respirator (N95 mask, elastomeric mask or PAPR) for a HIS/COVID-19 patient receiving an AGP. Five consultants (4%) report using only a surgical mask during AGPs for HIS/COVID-19 patients.

**Table 2:** Heat map of PPE chosen by EDSMOs for various clinical scenarios. Low (LIS) or high (HIS) index of suspicion for COVID-19. Aerosol generating procedure (AGP)

| PPE | Non-Patient Care | Tea Room | Toilet | LIS | HIS | HIS + AGP |
| --- | --- | --- | --- | --- | --- | --- |
| Face shield | 1% | 0% | 2% | 4% | 71% | 75% |
| Safety glasses/goggles | 1% | 0% | 1% | 12% | 79% | 76% |
| Surgical masks | 31% | 9% | 10% | 61% | 71% | 34% |
| Reusable fabric masks | 2% | 1% | 1% | 2% | 6% | 5% |
| N-95 masks/respirators | 0% | 0% | 1% | 6% | 48% | 83% |
| Elastomeric respirators | 0% | 0% | 0% | 0% | 3% | 2% |
| PAPR | 0% | 0% | 0% | 0% | 1% | 2% |
| Disposable surgical hat | 0% | 0% | 1% | 2% | 25% | 29% |
| Reusable Surgical hat | 0% | 0% | 1% | 4% | 7% | 7% |
| Standard disposable gown | 0% | 0% | 1% | 13% | 87% | 84% |
| Full-body impermeable suit | 0% | 0% | 0% | 2% | 6% | 7% |
| Gloves | 2% | 0% | 1% | 52% | 90% | 83% |
| Double gloves | 0% | 0% | 0% | 1% | 21% | 25% |
| Foot coverings | 0% | 0% | 0% | 1% | 16% | 13% |

PPE practice preferences vary when caring for either a High or Low Index of Suspicion patient while not performing an AGP. For a HIS/COVID-19 patient without an AGP consultants report N95 use of 48%, the rest using surgical mask alone or over N95. When seeing a LIS patient and no AGP, 6% report using an N95 respirator. Two-thirds (69%) wear some type of mask seeing LIS patients and one third of emergency consultants surveyed see LIS patients in their ED without a mask. While working outside of direct patient care but still in the hospital one third of ED SMOs wear a surgical or reusable fabric mask. Toilets may present a unique risk for droplet and possibly faecal-airborne transmission yet only 10% report using masks in toilets.(28, 29) A summary rank-ordered list by ED consultants’ assessment of their most likely source of exposure to COVID-19 identified “wearing inadequate PPE for patients not suspected of COVID-19 infection”, followed by “contracting it from fellow staff members” or “accidental doffing exposure” as the top three most likely routes of nosocomial infection. Consultants were less concerned about inadequate N95 mask fit testing or the lack of appropriate training or PPE for co-workers such as housekeeping staff (Table 3).

**Table 3:** Rank the most likely reason that you think puts you at risk of exposure to COVID-19 at work? (1 for most likely, 8 for least likely)

| Rank | Risk | Mean | C.I. |
| --- | --- | --- | --- |
| 1 | Wearing inadequate PPE for patient(s) not suspected of COVID-19 infection | 2.93 | 0.38 |
| 2 | Contracting it from a fellow staff member in the ED | 3.06 | 0.38 |
| 3 | Accidental PPE doffing exposure | 3.53 | 0.39 |
| 4 | Wearing inadequate PPE for patient(s) suspected of COVID-19 infection | 3.67 | 0.35 |
| 5 | Not being able to access adequate PPE | 4.44 | 0.48 |
| 6 | Inadequate mask fit testing for staff | 5.62 | 0.41 |
| 7 | Cleaners have been provided inadequate training and/or inadequate PPE | 5.66 | 0.32 |
| 8 | Not applicable- I do not fear risk of COVID-19 exposure at work | 6.57 | 0.57 |

## DISCUSSION

This study assesses the preparedness of EDs around Aotearoa New Zealand for the eventual reintroduction of SARS-CoV-2 (30). Survey results identify weaknesses in local NZ hospital infection control practices which have been cited as risks in prior outbreaks in other countries(7, 10, 15). Our results reveal incomplete ED engineering upgrades such that 12% of NZ emergency consultants report no access to negative flow rooms for AGPs, continued use of cohorted bed bays possibly collocates infected and non-infected patients in areas of shared air circulation, and crowded work environments are inconsistent with recommendations for physical distancing. Results also indicate variations in pandemic specific administrative policy, adherence and practice. In particular, only a third of specialists reported routine monitoring of donning and doffing of PPE. Only two-thirds would intubate a high index of suspicion (HIS) patient in a negative pressure room. High flow nasal cannula (HFNC) for hypoxic COVID-19 patients would be utilized by only half of ED consultants in this survey. Finally, infection control through PPE availability and proper use may be compromised by the finding that about one-tenth of ED consultants reported not being fit tested for N95 masks. Although reported N95 mask shortages were rare, only 87% of respondents would use a respirator in the high risk setting of a HIS/COVID-19 patient receiving an AGP.

When elimination of SARS-CoV-2 fails and adequate community-wide immunity has not been established it is these proven step-wise controls that are needed to curb nosocomial spread and prevent health-care system compromise. Cited as primary are engineering controls which decrease SARS-CoV-2 transmission by modifications to hospital ventilation, bed allocation, streaming patients and physical distancing of staff. A minimum requirement would provide enough adequately ventilated negative pressure rooms, or at least negative directional airflow, to allow for treatment of multiple respiratory isolation patients requiring AGPs. Negative flow dilutes contaminated air breathed by HCWs caring for patients with airborne transmissible infections. DHBs should prioritise ED patient areas with a greater number of room air changes per hour (ideally 6-12 ACH), and greater proportion of fresh (vs recycled) air or consider portable HEPA filter units if airflow is inadequate.(9, 20) The finding that 12% of consultants report no access to at least one negative flow room for AGPs, mostly in smaller peripheral hospitals, suggests NZ DHBs have not equitably upgraded all EDs.

Control of bed-allocation during a COVID-19 surge reiterates issues common to emergency systems chronically plagued by over-crowding and limited resources.(31) Somewhat unique to a respiratory pandemic, patients with suspected COVID-19 may compromise the capacity to protect other patients from exposure. Because of this, single rooms to isolate suspected cases or protect vulnerable non-infected patients become a premium. The delay between clinical suspicion and confirmatory testing can further prolong lengths of stay such that available, rapid SARS-CoV-2 testing must be a priority.(32) Our results show most NZ suspected COVID-19 patients are streamed to separate ED areas or wards away from others where possible. In the Australian model, HCWs are to be rostered such that they minimize intermingling between COVID and Non-COVID teams.(10, 20, 33) A majority (92%), of NZ consultants work in EDs that stream HIS/COVID-19 patients but most (64%) report being rostered to alternate seeing both HIS and LIS patients. Smaller EDs and limited rosters were often cited as the reason in the comments.

In some instances there may be pressure to cohort patients in multiple bed bays with shared air circulation. In this study, three quarters of NZ specialists report having ED patients cohorted with only curtains separating beds. Based on overseas experience, large numbers of COVID-19 patients in confined spaces may create a high density of aerosols and cause HCWs to stay longer as they attend each patient increasing their risk. Best practice reduces patient density to one per room (even if in a 2 or 4 bed bay) and mandates airborne PPE for staff in these situations.(10, 34, 35) Conversely, use of multi-bed bays to cohort presumed Non-COVID patients risks misidentifying the asymptomatic or presymptomatic patients as safe to collocate with other uninfected individuals(32). Well ventilated or HEPA filtered areas may decrease this risk but evidence is limited(36).

Although much attention is directed toward patient-to-HCW transmission, literature has identified HCW transmission to patients and to other HCWs and many of these nurses or doctors had no symptoms reiterating the importance of maintaining physical distancing and mask wearing in non-clinical areas when SARS-CoV-2 is circulating.(37, 38) Ranking this risk second in Table 2 suggests most NZ ED specialists may be aware of this concern. Despite recommendations to maintain physical distancing in non-clinical work areas most (86%) of NZ specialists disagreed that their ED workstations were engineered for adequate room (Figure 1). This illustrates how the lack of resources, physical space or personnel can undermine administrative efforts to protect staff and patients from exposures.

Administrative policy involves institution of rules that change how health care workers behave, it alters work flow and implements infection control protocols. Success may depend on dissemination of guidelines, staff confidence in recommendations, or practice (real life or simulation). This can be undermined by poor messaging, mistrust or when case counts are low and the risk no longer justifies the effort. Vaccination may also create a sense that these other controls are not needed.

Initial training for PPE use was universal (97%) but ongoing interval training was not common nor was mandatory observation during donning or doffing as recommended in the literature.(18) Training (baseline and refreshers) and monitoring policy for PPE use (spotters) for all clinical and non-clinical staff is not standardized across DHBs (Table 1). Simulations to practice skills (such as intubation and NIV use) and accommodate for PPE are variably applied in NZ(18).

Experience in other countries has shown HCW PPE breaches, exposures and infections cause large numbers of staff furloughs, worsening nurse to patient ratios and causing the remaining staff to experience high workloads.(10, 33, 39) Maintaining a healthy skilled workforce is paramount to offset predicted inadequate staffing. A proactive approach should be used to support infected and furloughed staff wellbeing, with dedicated nursing and medical staff monitoring physical and mental health and providing support. Given the gravity of HCW infection and the system failure it implies, every suspected healthcare associated infection should trigger a bundle of immediate infection control measures.(40)

Among the strongest recommendations in the literature regarding prevention of HCW nosocomial infection is to “decant” or decrease overcrowding of COVID-19 patients in EDs and wards.(10) Ensuring a manageable workload through adequate staffing ratios by anticipating the increased care required for these infectious respiratory failure patients is paramount. This may also prevent the added fatigue HCWs face secondary to PPE compliance, doffing observation, and decontamination of providers and work environment. These additional tasks are not being calculated into traditional bedside severity scores and underestimate nursing ratios.

Only two thirds of NZ ED specialists (64%) would intubate HIS/COVID-19 patients in a negative pressure/flow room. Allowing for the roughly 12% who reported no access to this engineering control, it suggests a quarter of NZ ED consultants would depart from recommended practice. It is possible the difference represents consultants who do not intubate and or intend to transfer care before that indication arises.

Personal Protective Equipment places a barrier between the HCW and the infectious agent (the principal example being respirators and other masks) and are considered the final and least effective control measure because it relies on consistent individual action at the point of care.(12) PPE should be implemented through clear guidelines and be current with peer reviewed literature and expert recommendations.(22, 41-43) The NZ Ministry of Health (MoH) last updated PPE recommendations 22 September 2020 and these do not promote use of N95 respirators outside of AGPs for HIS/COVID-19 patients.

This study shows that shortages of N95 masks, and one key reason for limiting their use, have largely resolved and improved supplies allow hospitals to stop contingency and crisis practices (e.g. decontaminating N95s and using surgical masks in place of respiratory protection). The scientific community has acknowledged literature demonstrating transmission through inhalation of small airborne particles is a significant mode of SARS-CoV-2 virus transmission.(40, 43-45) These studies demonstrate aerosols produced through breathing, talking, coughing and yelling can remain in air and viable for long periods of time, travel long distances within a room and sometimes farther depending on ventilation. The experience in The Royal Melbourne Hospital City Campus outbreak noted that “aerosol generating behaviour” (AGB) in infected patients appeared to be linked to transmission events(10). Patients shouting, vigorous coughing, cognitive impairment and combative behaviour, actions common in ED patients, should mandate airborne precautions.(39, 43)

Fit testing of N95 respirators in line with other nations’ health and safety legislation was late to be initiated in NZ, and for at least 15 consultants (11%) was still not available at the time of this survey.(18, 22) Small peripheral facilities, as was the case for negative flow rooms, appear to be less prepared.

In the scenario-based PPE questions (Table 2), the finding that up to 13% of NZ ED consultants would not choose an N95 respirator, elastomeric or PAPR in the context of an aerosol generating procedure (AGP) for a HIS/COVID-19 patient was unexpected and raises concern. Given the low prevalence of SARS-CoV-2 in NZ, the probability of an HIS patient being infected is low, but not zero. Some ED consultants may argue N95s are not necessary due to elimination efforts or may believe they are still in short supply. But the omission of this recommended PPE could be interpreted as a purposeful disregard of evidence based pandemic IPC practice or a deliberate ignorance of why these policies exist. In a pandemic, an individual’s choice to forgo personal protection does not just take the risk for themselves, but for the community of others on their health care team, the other patients they care for, and their families and close contacts. Instituting and maintaining a standardized observer system and breach protocols should remedy this issue and may help promote a culture of staff safety, risk and adverse event reporting and staff support. NZ has enjoyed near SARS-CoV-2-free medical practice but sporadic reintroduction has occurred with HCW infection and risking transmission during aerosol generating procedures is an unconscionable breach of infection prevention and control.(16) This will have to change as SARS-CoV-2 is reintroduced.

Some professional organizations have gone one step further and simplified the practice. The Australasian and New Zealand College of Anaesthesia in conjunction with the Infection Control Expert Group (ICEG) recently recommend wearing airborne precaution PPE for care of all patients with high risk of SARS-CoV-2, irrespective of the community transmission.(41)

## LIMITATIONS

Our study has several limitations. First, our electronic convenience sample of emergency consultants may not be representative of all ED consultants in NZ. Those motivated to respond may feel they have more or less access to protective policies than non-respondents. Second, we asked respondents to recall experiences or project how they would practice if they were caring for a COVID-19 patient. Although many consultants have carefully considered how they would provide care, the low case counts in NZ mean that many respondents have not provided that care. Finally, we adapted previously published inventories of COVID-19 protective practices. While these questions had not been specifically validated in our health system, we sought to ask about objective availability of specific countermeasures that could be expected to be measured accurately using direct questions.

## CONCLUSION

These survey results from NZ ED consultants identify potential risks of failure in the hierarchy of infection controls currently in place to prevent nosocomial spread of SARS-CoV-2 or future emerging infections. Our findings show that engineering upgrades to respiratory pandemic standards are not prevalent, administrative COVID-19 policy has not adapted to scientific advances seen in policy from other healthcare systems (ie. Australia), and PPE current practice reveals high variability suggesting poor dissemination of guidelines, low confidence in recommendations, or little practice because of low prevalence. NZ’s public health success in SARS-CoV-2 elimination and the promise of protective immunity through vaccines has allowed for a relaxation of other layers of infection control even as evidence-based practice supporting them has evolved. As New Zealand borders reopen and crowded and under resourced emergency departments and their frontline health care workers face endemic COVID-19, it appears possible to use lessons learned elsewhere to identify and better prepare for this phase of the pandemic.

## Data Availability

The Survey in PDF format will be attached as supplementary material online. Excel spreadsheet of results will be made available as supplementary material. There are unpublished concurrent studies using this data set at present.

## ACKNOWLEDGMENTS

We thank our many colleagues who have contributed to development of the NZEDC19 Preparedness Survey. In particular Drs. Arvind Rajamani, Ashwin Subramaniam, Jason Barton, Eugene Fayerberg, David Hammer, Elspeth Frascatore. Dr. Charlotte Chambers is employed as a research scientist with ASMS. ASMS administration did not have any role in the planning, writing or publication of the work or the decision to publish. We have no relevant disclosures or competing interests.

## Notes

### Competing Interest Statement

The authors have declared no competing interest.

### Funding Statement

No external funding was received

### Author Declarations

1) A waiver of ethical approval was received from Health and Disability Ethics Committee from the New Zealand Ministry of Health Courtney Parnell Administrator Health and Disability Ethics Committees 2)This work needs neither ethical approval nor locality assessment. Dr Michael Roberts Chief Medical Officer Northland District Health Board

### Summary of Updates

Reformatting of Tables and Figures. Editing length through condensed Results and Discussion

