## Supplemental Survey Questions for "New Zealand Emergency Department COVID-19 Preparedness Survey" (from another version of this preprint: version 1, 2021-04-07)

### Introduction

**The NZED COVID-19 Preparedness Survey focuses on your safety and well-being during the COVID-19 pandemic.**

**The questions are peer-reviewed and created with the help of emergency specialists and infectious disease experts focused on health care worker safety and well-being.**

**We hope the results from this research will enable dialogue as to best infection control measures, and assist with standardising local protocols with the aim of minimising risk of nosocomial COVID-19 infections in the Emergency Department (ED).**

**ASMS understands that your commitment to the welfare of your patients and colleagues is predicated on your ability to focus on providing the best medical care in the safest possible work environment. We understand this requires practice and preparation.**

**Please take this opportunity to relate the current and proposed practices in your ED and share your opinions and thoughts.**

**If you have any questions regarding this research, please do not hesitate to contact Dr Charlotte Chambers at ASMS:**

**Thank you for your time.**

### Emergency Department (ED) Facility Characteristics

1. **Where is your primary place of work?**

2. **Do you have any negative pressure <u>beds</u> in your ED and if so, how many?**

For the purposes of this survey, a negative pressure or negative flow bed is defined as any bed in single or multiple rooms with minimum of 6 air changes per hour with or without an ante room.

- We don't have any negative pressure beds in our ED
- 1-4
- 5-9
- 10-14
- 15-19
- 20 or more
- Other (please specify)

3. **How many beds in your ED at the time of this survey are in shared rooms for cohorted patients**

For the purposes of this survey shared rooms are defined as large rooms with multiple curtained beds

- 1-4
- 5-9
- 10-14
- 15 or more
- I'm not sure
- None
- Other (please specify)

4. **When you have either confirmed COVID-19 patients or high index of suspicion (HIS) patients entering your ED do you treat them in a separate area (separate flow/segregated ED)?**

- ◯ Yes
- ◯ No
- ◯ Not applicable
- ◯ I'm not sure
- ◯ Other (please specify)

5. **Some EDs may be segregated into COVID and Non-COVID areas for patient care. How would you expect to be personally rostered in your ED?**

- ◯ We don't have a segregated ED
- ◯ I will be treating COVID patients only
- ◯ I will be treating Non-COVID patients only
- ◯ I will see both, alternating between them as needed
- ◯ I'm not sure
- ◯ Comment:

6. **Do you feel able to meet minimum physical distancing requirements in the non-clinical staff areas of your ED?**

| | Strongly agree | Agree | Neither agree nor disagree | Disagree | Strongly disagree | Not applicable |
|---|---|---|---|---|---|---|
| Offices | ◯ | ◯ | ◯ | ◯ | ◯ | ◯ |
| Work stations on ED floor | ◯ | ◯ | ◯ | ◯ | ◯ | ◯ |
| Break rooms | ◯ | ◯ | ◯ | ◯ | ◯ | ◯ |
| Bathrooms | ◯ | ◯ | ◯ | ◯ | ◯ | ◯ |
| Changing rooms | ◯ | ◯ | ◯ | ◯ | ◯ | ◯ |
| Canteens/Cafeteria | ◯ | ◯ | ◯ | ◯ | ◯ | ◯ |

Comments:

### PPE training and fit testing

1. **What type of training have you had in the use of PPE at your current place of employment?**

Please select all that apply from the following:

- [ ] None - I have not completed any online training or employer required/directed training
- [ ] Self-taught using online training (video, reading material)
- [ ] In-person group demonstration in which I only watched
- [ ] In-person group session in which I practiced putting PPE on and removing it properly
- [ ] In-person individual demonstration in which I only watched
- [ ] In-person individual session in which I was observed putting PPE on and removing it properly
- [ ] Other (please specify)

2. **Approximately, how frequently are you trained in any of the following activities?**

| | Upon request | Never | Once | Annually | I'm not sure |
|---|---|---|---|---|---|
| Training for donning and doffing PPE | ○ | ○ | ○ | ○ | ○ |
| Simulation sessions on intubating COVID patients | ○ | ○ | ○ | ○ | ○ |
| Simulation session for non-invasive ventilation (NIV: CPAP, HFNC) | ○ | ○ | ○ | ○ | ○ |
| Simulation sessions on awake/self prone positioning of COVID patients | ○ | ○ | ○ | ○ | ○ |
| Simulation sessions on transporting COVID patients | ○ | ○ | ○ | ○ | ○ |

Other (please specify)

3. If you have been fit tested with a N-95 mask/respirator within the last 12 months, what method was used to determine fit?

- ○ Not applicable; I have not been fit tested within the last 12 months
- ○ Qualitative (odour or taste detection in hood)
- ○ Quantitative (machine sampling via tubing)
- ○ I'm not sure
- ○ Other (please specify)

### PPE Scenarios

1. **What personal precautions are you currently using in the following settings?**

Please select all that apply from the following:

| | When in your ED but not providing patient care (charting, making telephone calls) | When in your Break/Tea Room (eating, conversing with colleagues) | When in your ED providing care for a non-COVID-19 low index of suspicion (LIS) patient | When in your ED providing care for COVID-19 HIS or confirmed patient | When within 2m of an aerosol-generating procedure for a confirmed or HIS COVID-19 case | When using the bathroom facilities |
|---|---|---|---|---|---|---|
| Standard precautions (handwashing, distancing from others) | ☐ | ☐ | ☐ | ☐ | ☐ | ☐ |
| Face shield | ☐ | ☐ | ☐ | ☐ | ☐ | ☐ |
| Safety glasses/goggles | ☐ | ☐ | ☐ | ☐ | ☐ | ☐ |
| Surgical masks | ☐ | ☐ | ☐ | ☐ | ☐ | ☐ |
| Reusable fabric masks | ☐ | ☐ | ☐ | ☐ | ☐ | ☐ |
| N-95 masks/respirators | ☐ | ☐ | ☐ | ☐ | ☐ | ☐ |
| Elastomeric respirators | ☐ | ☐ | ☐ | ☐ | ☐ | ☐ |
| Powered air-purifying respirator systems (PAPR CAPR) | ☐ | ☐ | ☐ | ☐ | ☐ | ☐ |
| Disposable surgical hat | ☐ | ☐ | ☐ | ☐ | ☐ | ☐ |
| Reusable surgical hat | ☐ | ☐ | ☐ | ☐ | ☐ | ☐ |
| Standard disposable isolation gown | ☐ | ☐ | ☐ | ☐ | ☐ | ☐ |
| Full-body impermeable suit | ☐ | ☐ | ☐ | ☐ | ☐ | ☐ |
| Gloves | ☐ | ☐ | ☐ | ☐ | ☐ | ☐ |
| Double gloves | ☐ | ☐ | ☐ | ☐ | ☐ | ☐ |
| Foot coverings | ☐ | ☐ | ☐ | ☐ | ☐ | ☐ |

Other (please specify):

____________________

2. **Does your ED have a formal policy on showering/shampooing after caring for COVID-19 patients?**

| | Mandatory | Not mandatory | Not practical as we have limited facilities | We don't have shower facilities | Not aware of a formal policy |
|---|---|---|---|---|---|
| Immediately after every single patient-contact episode | ◯ | ◯ | ◯ | ◯ | ◯ |
| Only if PPE was breached | ◯ | ◯ | ◯ | ◯ | ◯ |
| At the end of the shift | ◯ | ◯ | ◯ | ◯ | ◯ |
| After reaching home | ◯ | ◯ | ◯ | ◯ | ◯ |

Comment:

3. **In your ED, is PPE donning monitored by an observer prior to care for COVID-19 patients?**

◯ Yes, all the time (mandatory)

◯ Yes, some of the time (ad-hoc)

◯ No

◯ I'm not sure

◯ Other (please specify)

4. **In your ED, is PPE doffing (removal) monitored by an observer to identify breaks in doffing technique after care for COVID-19 patients?**

◯ Yes, all of the time (mandatory)

◯ Yes, some of the time (ad-hoc)

◯ No

◯ I'm not sure

◯ Other (please specify)

### Policies and practices for patients with suspected COVID-19

1. **For endotracheal intubation, which of the following (if any) is your ED using all or most of the time for patients with:**

Please select all that apply from the following:

| | Low Index of Suspicion for COVID-19 | High Index of Suspicion/Confirmed COVID-19 |
|---|---|---|
| Negative pressure bed (bed with minimum of 6 air changes per hour) | ☐ | ☐ |
| Video laryngoscopy | ☐ | ☐ |
| Intubation barrier protection (e.g., intubating boxes , intubating bags, etc.) | ☐ | ☐ |
| Intubation barrier protection with integrated suction (e.g., intubating boxes connected to suction; negative pressure hood) | ☐ | ☐ |
| Intubation response teams (with dedicated staff) | ☐ | ☐ |
| Intubation through a supraglottic device (e.g., intubating LMA, etc.) | ☐ | ☐ |
| None of these | ☐ | ☐ |

Other (please specify)

2. **Does your ED have a dedicated intubation team for confirmed or suspected COVID-19 patients?**

- ◯ Yes, consisting of ICU/Anaesthesia responding to ED
- ◯ Yes, consisting of ICU/ED formalised
- ◯ Yes, ED only
- ◯ No
- ◯ I'm not sure
- ◯ Other (please specify)

3. **For patients with confirmed or suspected COVID-19, is your ED practice/protocol that patients will be treated with high-flow nasal cannula (HFNC), if needed?**

- ◯ Yes
- ◯ No
- ◯ I'm not sure
- ◯ We don't have a formal protocol
- ◯ Comment:

4. **For ED patients with confirmed or suspected COVID-19, is your ED practice/protocol that patients will be treated with non-invasive positive pressure ventilation (NIV, including CPAP or BiPAP), if needed?**

- ◯ Yes, in any area in the ED
- ◯ Yes, only in a negative flow/pressure room in ED
- ◯ Yes, only with in-line or expiration viral filter in negative flow/pressure room in ED
- ◯ Yes, only after transfer to the ICU if appropriate
- ◯ No
- ◯ We don't have a formal protocol
- ◯ Other (please specify)

5. **For confirmed or suspect COVID-19 ED patients, under what circumstances might NIV (including CPAP or BiPAP) be used in your ED?**

**Please select from the following:**

- ☐ Any patient with respiratory failure that I think will benefit from NIV if indicated (NIV: CPAP/BiPAP)
- ☐ Only patients that have other co-morbidities known to benefit from NIV (eg. COPD, CHF, OSA)
- ☐ Only patients who have a "Do Not Intubate" or a "Do Not Resuscitate" order
- ☐ Only when mechanical ventilators are scarce
- ☐ Other (please specify)

### PPE Breaches

1. **Please select from the following scenarios what constitutes a 'Breach in PPE' in your Emergency Department**

| | Breach | Not a breach | Not aware of formal policy |
|---|---|---|---|
| Inadequate face protection | ○ | ○ | ○ |
| Inadequate eye protection | ○ | ○ | ○ |
| Improper donning/doffing | ○ | ○ | ○ |
| Exposure of skin due to a glove or gown tear | ○ | ○ | ○ |
| Inadequate shoe cover | ○ | ○ | ○ |
| Direct contact of skin to any secretion | ○ | ○ | ○ |
| Needle stick | ○ | ○ | ○ |
| Poor mask fit | ○ | ○ | ○ |

Other (please specify)

2. **What measures have been advised by your hospital/ED administration when a PPE breach has been identified?**

| | Mandatory | Optional | Not aware of a formal policy |
|---|---|---|---|
| Shower immediately | ○ | ○ | ○ |
| Report to ID/designated authorities | ○ | ○ | ○ |
| Retraining given for donning or doffing | ○ | ○ | ○ |
| Quarantine with testing protocol | ○ | ○ | ○ |

Other (please specify)

### PPE supply and re-use

1. At the time of this survey, **please select from the following any PPE that is <u>out of stock or otherwise unavailable for clinical use</u> in your ED:**

- [ ] Reusable face shields
- [ ] Disposable face-shields (single use)
- [ ] Safety glasses/goggles
- [ ] Surgical masks
- [ ] Reusable fabric masks
- [ ] N-95 masks/respirators
- [ ] Elastomeric respirators
- [ ] Powered air- purifying respirator systems (PAPR, CAPR, etc.)
- [ ] Disposable surgical hat
- [ ] Reusable surgical hat
- [ ] Standard disposable isolation gown
- [ ] Full-body impermeable suit
- [ ] Gloves
- [ ] Foot coverings
- [ ] Other (please specify)

2. **Have you ever re-used PPE equipment according to any of the following scenarios?**

| | Yes | No | I'm not sure |
|---|---|---|---|
| We re-use N-95 masks without sterilization | ○ | ○ | ○ |
| We re-use N-95 masks after sterilization | ○ | ○ | ○ |
| We re-use face shields without washing with cleaning solution | ○ | ○ | ○ |
| We re-use face shields after washing with cleaning solution provided | ○ | ○ | ○ |
| We re-use goggles without washing with cleaning solution | ○ | ○ | ○ |
| We re-use goggles after washing with cleaning solution provided | ○ | ○ | ○ |

Other (please specify)

### Your views

1. **Which of the following best describes your level of confidence in your ED's PPE protocol?**

| | **During Level 4 Lockdown** | **At the time of this survey** |
|---|---|---|
| I am/was confident that our PPE protocol will keep me completely safe | ☐ | ☐ |
| I think my ED's protocol put me at risk and that I should have better PPE than is available, or use PPE more often than required by protocol | ☐ | ☐ |
| I think my ED's PPE protocol is too restrictive, and I feel that I can safely practice without wearing PPE every time that it is required by protocol | ☐ | ☐ |
| I am/was unsure about the safety of our PPE protocol and feel neither safe or unsafe | ☐ | ☐ |
| I am/was not aware of a PPE protocol in my ED | ☐ | ☐ |

Other (please specify)

2. **At the time of the initial COVID-19 outbreak in New Zealand (during level 4 lockdown), please consider how you felt about the following:**

| | Never | Rarely | Sometimes | Often | Always |
|---|---|---|---|---|---|
| I worried that family members or other close contacts were at risk of exposure to COVID-19 because of my work | ○ | ○ | ○ | ○ | ○ |
| I worried that I was at risk of exposure to COVID-19 because of my work | ○ | ○ | ○ | ○ | ○ |
| I worried about the level of preparedness of my hospital and ED | ○ | ○ | ○ | ○ | ○ |
| I worried about the supply of adequate and appropriate PPE in my ED | ○ | ○ | ○ | ○ | ○ |
| I felt anxious and stressed because of COVID-19 | ○ | ○ | ○ | ○ | ○ |

Other (please specify)

3. **Please answer the following questions regarding your views on how prepared your ED would be if there was another wave of COVID-19 in New Zealand**

| | Strongly agree | Agree | Neither agree nor disagree | Disagree | Strongly disagree |
|---|---|---|---|---|---|
| Our ED is prepared and ready for another wave of COVID-19 | ☐ | ☐ | ☐ | ☐ | ☐ |
| Our ED would have adequate staffing levels if there was another wave of COVID-19 | ☐ | ☐ | ☐ | ☐ | ☐ |
| Our ED would have adequate supplies of appropriate PPE if there was another wave of COVID-19 | ☐ | ☐ | ☐ | ☐ | ☐ |
| We would be able to rapidly test and in a timely manner diagnose possible cases of COVID-19 | ☐ | ☐ | ☐ | ☐ | ☐ |

Other (please specify)

____________________

4. **Do you feel you are at risk of infection from COVID-19 at work?**

| Strongly agree | Agree | Neither agree nor disagree | Disagree | Strongly disagree |
|---|---|---|---|---|
| ○ | ○ | ○ | ○ | ○ |

Other (please specify)

____________________

5. **If you feel you are at risk of COVID-19 infection AT WORK, please rank the most likely reason that you think would put you at risk of exposure to COVID-19?**

**Please select 1 for the most likely reason through to 8 as the least likely.**

- [ ] Not applicable- I do not fear risk of COVID-19 exposure at work
- [ ] Not being able to access adequate PPE
- [ ] Wearing inadequate PPE for patient(s) not suspected of COVID-19 infection
- [ ] Wearing inadequate PPE for patient(s) suspected of COVID-19 infection
- [ ] Contracting it from a fellow staff member in the ED
- [ ] Accidental PPE doffing exposure
- [ ] Cleaners have been provided inadequate training and/or inadequate PPE
- [ ] Inadequate mask fit testing for staff

6. We would be grateful to hear your thoughts on any other aspects regarding the level of preparedness of your ED or the impact of COVID-19
